# Quantifying the impact of individual and collective compliance with infection control measures for ethical public health policy

**DOI:** 10.1101/2021.12.02.21267207

**Authors:** Daniel Roberts, Euzebiusz Jamrozik, George S. Heriot, Anja C. Slim, Michael J. Selgelid, Joel C. Miller

## Abstract

Infectious disease control measures often require collective compliance of large numbers of individuals to benefit public health. This raises ethical questions regarding the value of the public health benefit created by individual and collective compliance. Answering these requires estimating the extent to which individual actions prevent infection of others. We develop mathematical techniques enabling quantification of the impacts of individuals or groups complying with three public health measures: border quarantine, isolation of infected individuals, and prevention via vaccination/prophylaxis. The results suggest that (i) these interventions exhibit synergy: they become more effective on a per-individual basis as compliance increases and (ii) There is often significant “overdetermination” of transmission: if a susceptible person contacts multiple infectious individuals, an intervention preventing one transmission may not change the ultimate outcome (thus risk imposed by some individuals may erode the benefits of others’ compliance). These results have implications for public health policy during epidemics.

## Introduction

Public health policies for infectious diseases often require collective action. Among other things, this is because the actions of one individual can impact whether others are exposed to infection. Further, the protective impact of one individual’s actions on others may be augmented or undermined by the actions of others.

Increased population compliance with effective measures against infectious disease can lead to larger health benefits. However, few studies have examined the relationship between individual compliance and public health benefits. Yet ethical assessments of community infection control measures (especially those that are mandated for individuals) arguably depend in part on the extent to which individual actions contribute to collective benefits. There has, to date, been relatively little mathematically-informed *ethical analysis* of public health policy (*1, 2*).

According to established principles of public health ethics, policies are more ethically justifiable to the extent that they are expected to produce net public health benefits (outweighing the burdens or harms of the interventions) (*3*–*5*) Further, policies that involve limitations on individual freedom might be more justifiable to the extent that the behavior restricted by policy is likely to result in harm to others (*5*). It can also be ethically relevant to consider the causal pathways by which harm results (e.g. due to infectious disease transmission), in part because more complex causal pathways might have implications for the extent to which individual actions prevent harm that otherwise would not have occurred (*6*). Thorough assessments of these ethical considerations arguably require a quantitative methodology that estimates the (beneficial or harmful) impact of the actions of a single individual.

Significant mathematical modeling efforts have focused on health and economic impacts of SARS, pandemic influenza, and COVID-19 as well as the impacts of public health interventions (*7, 8*). However, although numerous ethical considerations are directly relevant to the justification of epidemic control policies (*9, 10*) investigations of the ethical implications of pandemic response in general and the COVID-19 response in particular have typically focused on allocation of scarce resources (*9, 11*–*14*), disparities in health outcomes (*15*), and issues of research ethics (*16, 17*), as opposed to quantitatively-informed ethical analysis of the benefits and harms of control policies and individual compliance with public health measures.

This article examines mathematical modeling techniques we have developed to explore how individual and collective behavior changes can affect two specific outcomes:

- The probability an epidemic becomes established in a population.
- The total number of infections that occur once an epidemic is established.

Our modeling approaches allow us to quantify the impact of a single individual’s behavior on these population outcomes. The models are designed to measure the impact of ethically salient aspects of transmission dynamics, including overdetermination and superspreading.

- **Overdetermination** occurs when a given outcome has more than one sufficient cause. For example, whether (i) an epidemic becomes established in a population or whether (ii) a specific individual becomes infected in an epidemic may be “overdetermined” when, respectively, (i) there are multiple introductions into a population (each of which would have been sufficient to cause an epidemic) and (ii) an individual is exposed to multiple infectious people (where each exposure would have been sufficient to infect the individual in question). This might be ethically salient because where overdetermination is significant (*e*.*g*., where there are multiple introductions into a population or in high transmission settings where each susceptible person experiences multiple exposures to infection), one (potentially) infectious person changing their behavior to reduce their risk of infecting others might make less difference to harmful outcomes because these outcomes will be more likely to occur in any case, due to the risk imposition of others.
- **Superspreading** diseases, including COVID-19, are characterized by the tendency for a small fraction of infected individuals to cause a large proportion of all transmissions, while most cause few or even no transmissions (*18*–*22*). Among other things, this affects the probability that a single introduction leads to established transmission in the population (*22, 23*). Because most individuals cause very little transmission, the disease typically only becomes established if there is an early superspreading event. We therefore explore the impact of superspreading (or ‘dispersion’ of the offspring distribution) by comparing the expected spread of an epidemic where superspreading is uncommon with an epidemic where the average number of transmissions is the same, but superspreading accounts for a high proportion of the transmissions.

Our primary goal is to examine the development and application of mathematical models to investigate the impact of individuals’ compliance with infectious disease control measures. We measure the effectiveness of relevant behaviors in terms of their impact on (i) the probability of an epidemic occurring (i.e., a chain of transmission begins and spreads widely in the population) and (ii) the total number of infections caused in an epidemic (we assume that morbidity and mortality and other costs are directly proportional to the number of infections, so total infections is a reasonable proxy of impact). These are both affected by overdetermination and superspreading, both of which are closely connected to random (stochastic) events. The methods we develop allow us to understand how stochasticity influences outcomes. The specific behaviors we investigate are isolating to prevent an epidemic from starting, and—if an epidemic is established—behavior changes to avoid infection or onwards transmission.

### The model

We briefly describe the model and define some terminology. More detail is in the Methods section. Our focus is on the impact of a decision made by an individual or a group to adopt some control measure, in a very large population. Prior to this decision, we assume that some background control measures are in place. We assume throughout that these background control measures will be maintained throughout the epidemic, and we are analyzing the impact of additional behavior changes.

The *offspring* of an infected individual are those who are directly infected by the individual. The *descendants* are those who are infected through a chain of transmissions starting at the individual.

The *offspring distribution* is the distribution of the number of transmissions infected individuals cause before recovery under whatever background control measures are in place. The variation in individual infectiousness is captured through choosing the appropriate distribution. We assume that the distribution does not change, except that once an individual has been infected, receiving additional transmissions has no impact (immunity following infection is complete). The average of the offspring distribution is the *reproduction number under control* R _*c*_. The population is assumed to be very large such that a large number of individuals must change behavior to have any impact on the average of the offspring distribution.

We study two offspring distributions, a *Poisson* distribution for which superspreading does not play an important role, and a *negative binomial* distribution for which superspreading is significant.

The *probability* of an epidemic P is the probability that the introduced disease does not go extinct at early times. This depends on the offspring distribution. If an epidemic occurs, it grows until depletion of the susceptible population limits its spread. The *attack rate* A is the fraction of the population infected in an epidemic. In a population with homogeneous susceptibility, A depends on the average of the offspring distribution R _*c*_ (*23, 24*).

In the remainder of this paper, we begin by describing the major results of our model, showing that individual actions produce benefits, in particular by avoiding transmission of infection to others. We find evidence for a synergistic effect: as more individuals comply with an intervention, the per-individual population benefit is increased. We also find evidence for the inverse effect: the potential benefits of some individual actions are undermined by the risk imposition of others, especially at high levels of transmission when overdetermination of infection is more common. We end the paper with a description of the mathematical tools that we have developed which allow us to quantify the benefit from behavior change at the individual scale.

We began this analysis in 2019, focusing on the role of Measles vaccine mandates and contact tracing, but with a goal of understanding the broader implications of public health measures and individual actions on infectious disease. Thus, this paper is intended to develop ideas and mathematical methods for a broad range of diseases. Our results have implications for newly emerging diseases such as SARS-CoV-2, Monkeypox, Ebola, or Zika, as well as for most childhood vaccine-preventable diseases. Because of the obvious context of SARS-CoV-2, we address nuances related to applying our results to SARS-CoV-2 in the discussion section (e.g., how immune-evading variants affect the applicability of our results).

## Results

We focus on the impact of three behaviors:

- *Border Quarantine*: Changing the behavior of individuals arriving to a population to prevent an epidemic from occurring
- *Case isolation*: Changing the behavior of newly infected individuals during an epidemic to prevent transmission to others.
- *Vaccination or Prophylaxis*: In an ongoing epidemic, some individuals may take extra actions such as vaccination or prophylaxis to reduce their own risk of becoming infected and thereby reduce their risk of transmitting to others.

As a general rule, we find that the per-individual impact of multiple individuals changing behavior generally increases as more individuals change behavior. Motivated by this, for each of these three interventions we address three questions:

- What is the expected impact if a single individual adopts the behavior while everyone else continues as normal?
- What is the average impact per individual if a certain fraction of the population adopts the behavior?
- What is the marginal impact if one more individual were to adopt (or abandon) the behavior after a fraction has adopted it?

In all cases we consider how (or if) results are affected by the choice of offspring distribution, comparing how Poisson distributions compare with negative binomial distributions having the same mean.

### Border Quarantine

Many communities have historically had restrictions to prevent the introduction of individuals with infections from one geographic area or (sub-)population to another. Recently quarantines have been widely used in response to COVID-19. If an infected individual enters the population either by evading quarantine or due to ineffective quarantine, there is a chance that an epidemic may result (that is the outbreak becomes established and grows until depletion of susceptibles prevents its spread). However, by random chance, the infected individual perhaps would not cause any transmissions or only start a small chain of infections that quickly dies out. Conversely, if multiple introductions occur, averting transmission from any one infected individual may not prevent an epidemic (i.e., the outcome of an epidemic is overdetermined).

We start by analyzing how an individual’s isolation impacts the probability of an epidemic. Mathematically, this requires some results of probability generating functions (*23*). Our analysis allows us to investigate the role of overdetermination on the establishment of an epidemic, and to introduce the framework by which we later evaluate individual actions to reduce epidemic spread.

To quantify the impact of quarantining new arrivals, we begin by considering the introduction of a single infected individual into a large completely susceptible population. Based on assumed knowledge about the offspring distribution we calculate the probability that this single introduction results in an epidemic (i.e., it does not go extinct until susceptible depletion becomes important). From this it is a straightforward calculation to look at what happens if multiple introductions occur.

Given a known offspring distribution, the probability P that a single introduction into a large completely susceptible population results in an epidemic can be calculated using probability generating functions (*22, 23*).

Figure 1 shows the probability that a single infected individual entering a completely susceptible well-mixed population would cause an epidemic, as a function of R _*c*_, for our two offspring distributions. For R_*c*_ <1 we find P = 0, while for R_*c*_ >1 we find P > 0. The value of P increases with R _*c*_ but decreases as the offspring distribution becomes more heterogeneous. It should be noted however, that in the case of higher heterogeneity, the early growth of those outbreaks that do become established epidemics is typically higher than expected on average because they are likely to be seeded by superspreading events (*23*).

**Figure 1:**
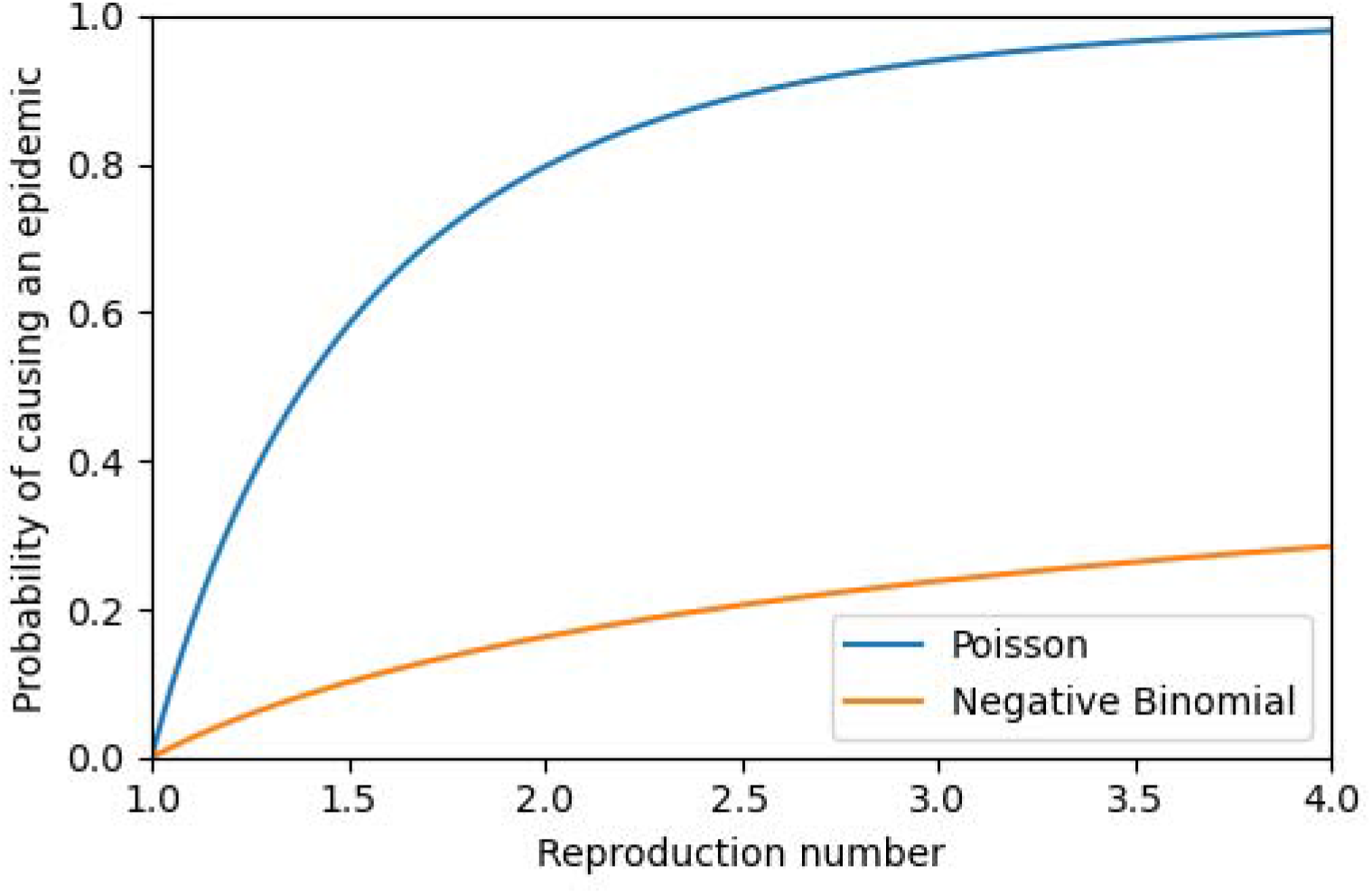
The probability a single infected individual entering a completely susceptible population causes an epidemic, assuming no other introductions. The offspring distribution follows either a Poisson distribution or a negative binomial distribution with a dispersion parameter of 0.16.

When there are multiple introduced infections, each may be separately sufficient to spark an epidemic (the establishment of an epidemic may be overdetermined). If there are multiple sufficient introductions, then isolating any one particular infected individual produces little to no public health benefit. The impact of a single isolating infectious individual on epidemic probability depends on the number of total infectious individuals introduced (*M*) and how many of them isolate (*L*) as shown in Figure 2. Overdetermination plays a large role in the trends in these figures. Each individual who does not isolate has an independent chance of triggering an epidemic. If even a single individual does trigger an epidemic, then those who isolate have no impact on the epidemic probability.

**Figure 2.**
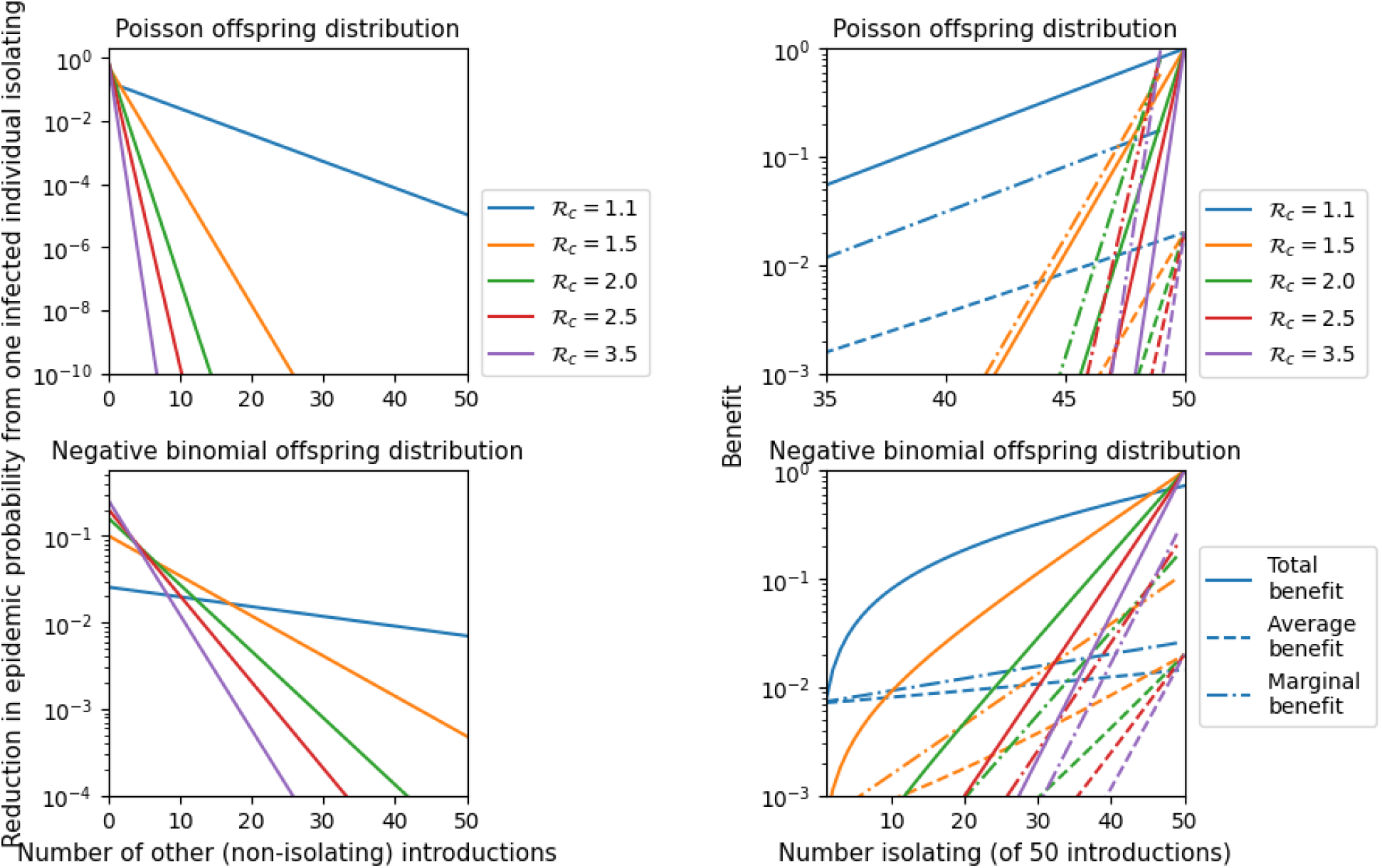
The benefit measured as the reduction in probability of an epidemic. We use a Poisson offspring distribution and a negative binomial offspring distribution with dispersion 0.16. We assume that L infected individuals enter the population of whom M isolate. **Left** (note different vertical scales): The reduction in epidemic probability if M = 1 (only one isolates) as a function of the number (L −1) who do not isolate. Due to overdetermination, this decreases as L increases. **Right** (note different horizontal scales): The reduction in probability if M isolate out of L = 50 introduced cases. This shows the total reduction in epidemic probability (solid line), the reduction averaged across the M isolating individuals (dashed line), and the marginal benefit of the M + 1 ^th^ isolating individual, (i.e., the change in probability from the next individual isolating) (dash-dot line). Each additional individual who isolates increases the average impact of all isolating individuals, as a consequence of declining overdetermination.

More specifically, if the probability that a single infected individual would trigger an epidemic is larger (larger R _*c*_), only a small number of non-isolating infected individuals are needed to effectively eliminate the impact of all those who isolate, shown in the left panels of Figure 2. Thus, border quarantine to prevent an epidemic is unlikely to be effective unless almost all infected individuals isolate (it should be noted that although it is a low probability event the societal benefit would be large). Since almost all infected individuals must isolate, in the absence of an effective screening test the quarantine must apply broadly, significantly increasing the costs and burden.

As overdetermination declines with greater compliance, we find evidence of synergy: the per-individual effectiveness of those who do isolate is increased as the number that isolate increases. Each additional isolating infected individual increases the effectiveness of the others who quarantine. The per-isolating individual impact grows with increasing numbers of infected individuals isolating. We see this in the right panels of Figure 2 where the average benefit per isolating infected individual increases as more isolate.

If even a few infected individuals fail to abide by border quarantine, however, this may significantly undermine the efforts of those who quarantine. As greater numbers of individuals comply, the benefits of an additional individual’s compliance and the population-level risks attached to one’s noncompliance both become greater.

### Public health measures during an epidemic

We now analyze the impact of behavior changes on the total number of infections in a population assuming an epidemic is established. We consider two different actions an individual could take:

- Isolation after infection to avoid transmitting further (such as isolation after a rapid test) which does not affect one’s own probability of becoming infected but does protect others, or
- Preventative actions taken in advance to avoid infection and hence also onwards transmission (such as vaccine or prophylactic medication).

### Isolation following infection

We begin here focusing on the mathematically simpler situation, isolation of a newly infected individual. As before, we look first at the effect of a single individual and then the effect of a collective behavior change. We assume the control measures remain constant for the duration of the epidemic.

We now consider the expected (average) number of averted infections from a single individual isolating *after* she is exposed (and develops infection), but before she becomes infectious. So we assume that her probability of infection is unaffected, but that if compliant with effective isolation measures she causes 0 additional infections. This represents the ideal situation and addresses the benefit from identifying a single individual prior to her infectious period. Clearly if an individual is identified after onset of infectiousness or only partly isolates, the expected benefit will be smaller.

We denote the expected total number of averted infections from a single newly infected individual isolating by F (R_*c*_). We account for overdetermination when calculating F (R_*c*_). That is, in calculating F (R_*c*_) we exclude those who would be infected through another transmission chain (overdetermination), but otherwise consider all “descendants”, i.e., cases linked by a chain of transmission from an infected individual. For R_*c*_ >1, overdetermination plays a role and the impact of overdetermination grows with R _*c*_. For R_*c*_ ≤1, overdetermination is negligible (for large well-mixed populations). Our results show that the value of F is independent of the details of the offspring distribution, it depends only on the average, R _*c*_. It turns out [see methods for derivation] that if R _*c*_ < 1 then 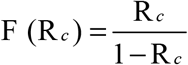. However if R _*c*_ > 1, then 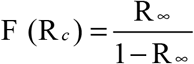 where R_∞_ = (1− A)R _*c*_ is the effective reproduction number at the end of the epidemic (equal to the final proportion susceptible 1− A times the reproduction number R _*c*_).

Figure 3 illustrates that close to R_*c*_ =1 the value of F (R_*c*_) is very large, approaching infinity in successfully isolates and avoids causing any infections is very large if R _*c*_ is close to 1. However, it becomes quite small if R _*c*_ is either large (because of overdetermination) or small (because there is little transmission). In practical terms, this analysis will be valid for large or small R _*c*_, but when R _*c*_ is close to 1, this analysis will break down if F (R_*c*_) is comparable to the population size. If F (R_*c*_) is less than 1% of the population size, we expect this to approximate the average number of averted infections per isolation. In any finite population, the prediction close to R _*c*_ = 1 is not attainable. Eventually a large enough fraction of the population is infected that independence assumptions in the analysis break down, and so the true number of averted infections will be smaller. Additionally, the transmission chains near R _*c*_ = 1 are very long and so results near this limit will also be more affected by seasonal changes or time-varying interventions.

**Figure 3.**
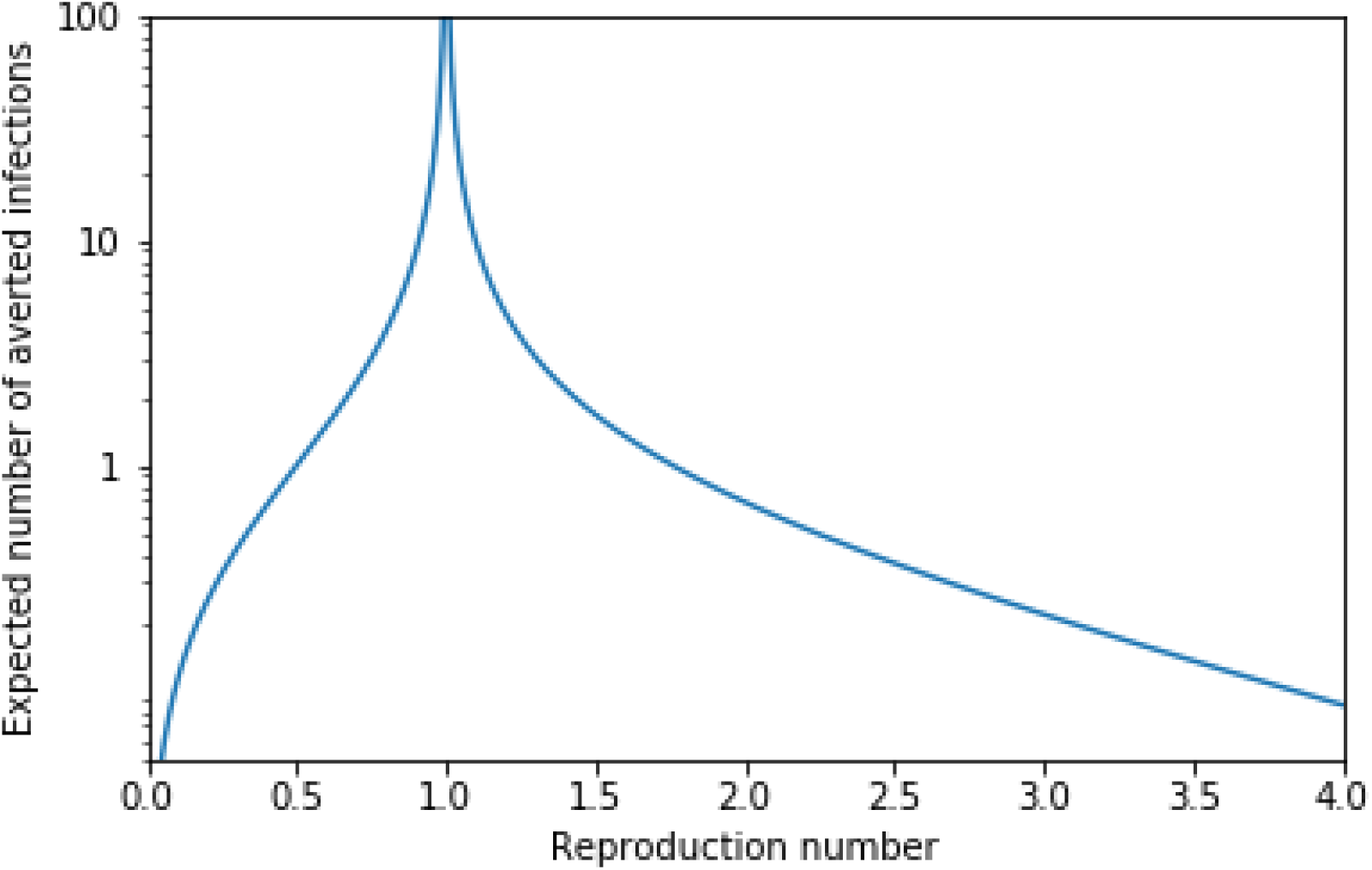
The expected number of averted infections, F (R_c_), due to a single infected individual isolating after infection and not transmitting, after accounting for overdetermination. The result is independent of the specific offspring distribution. This depends only on the average, R _c_. Note the divergence to ∞ as the reproduction number R_c_ →1, and the decay to 0 as R _c_ decreases towards 0 or increases towards ∞.

In addition to the *expected number* of infections averted, F (R_*c*_), it is possible to calculate the *distribution* of the number of infections averted. This is shown in Figure 4 for the Poisson and negative binomial distributions. Although the averages F (R_*c*_) are the same, the distributions are different. In the negative binomial case (and more generally for heavier tailed distributions) it is more common that either 0 infections are averted or a large number of infections are averted as compared to the Poisson case with the same R _*c*_. As is often seen in systems at a critical threshold, when R_*c*_ =1 the distributions are given by powerlaws. This figure shows that although it is rare for the number of averted infections to be large, the large events are frequent enough to produce a large average, F (R_*c*_), near R_*c*_ =1.

**Figure 4:**
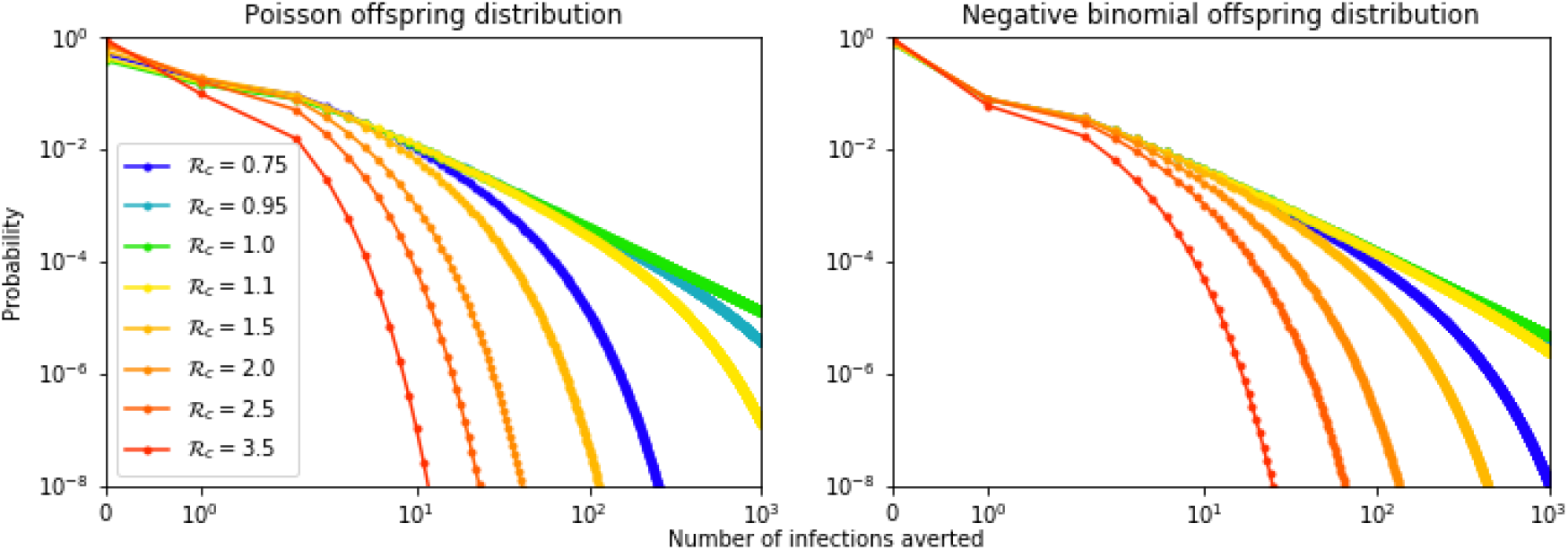
The distribution of the number of if a single individual isolates after infection. Calculated for Poisson and negative binomial distribution (with dispersion parameter 0.16). For R_c_ =1 the result is a power law distribution. If R _c_ is larger or smaller than 1, the distribution falls off quicker. For a given R _c_, both distributions have the same mean, but the negative binomial distribution results in a more heterogeneous outcome (both 0 infections averted or many infections averted become more common in distributions with a heavier tail).

If the individual does not isolate immediately and so is infectious for a short time, then obviously the expected benefit is smaller. If the expected number of transmissions is reduced to *ϕ*R _*c*_ 0 < *ϕ* <1, then the expected number of infections averted is (1−*ϕ*)F (R_*c*_).

Having analyzed the impact of a single individual who isolates following infection, let us now consider what happens if some nonzero fraction of the infected individuals isolate. If the number is not large enough to materially affect R_*c*_, then each isolation is effectively independent, and to find the expected benefit, we can simply multiply F (R_*c*_) by the number who isolate. However, if a nonvanishing fraction isolate this will alter R_*c*_. Then we see an increasing marginal benefit of collective compliance until R_*c*_ reaches 1. Each additional individual who isolates increases the effectiveness of those who have already isolated by reducing the level of overdetermination.

Figure 5 shows a similar trend to Figure 2, with a synergistic effect: if a larger fraction of infected individuals isolate, the expected number of infections averted per isolating individual grows. So, their combined impact grows faster than linearly in the fraction isolating. If R _*c*_ is large and very few infected individuals are isolating, then those who do have little impact. However as the fraction of infectious individuals who isolate increases, the average impact grows. As the system nears the epidemic threshold the average impact grows large, and the impact of each additional infected individual isolating diverges. Past the threshold where epidemics become impossible, the average impact is infinite (in the large-population limit) because only a small number of individuals isolate, but in doing so they eliminate the epidemic.

**Figure 5:**
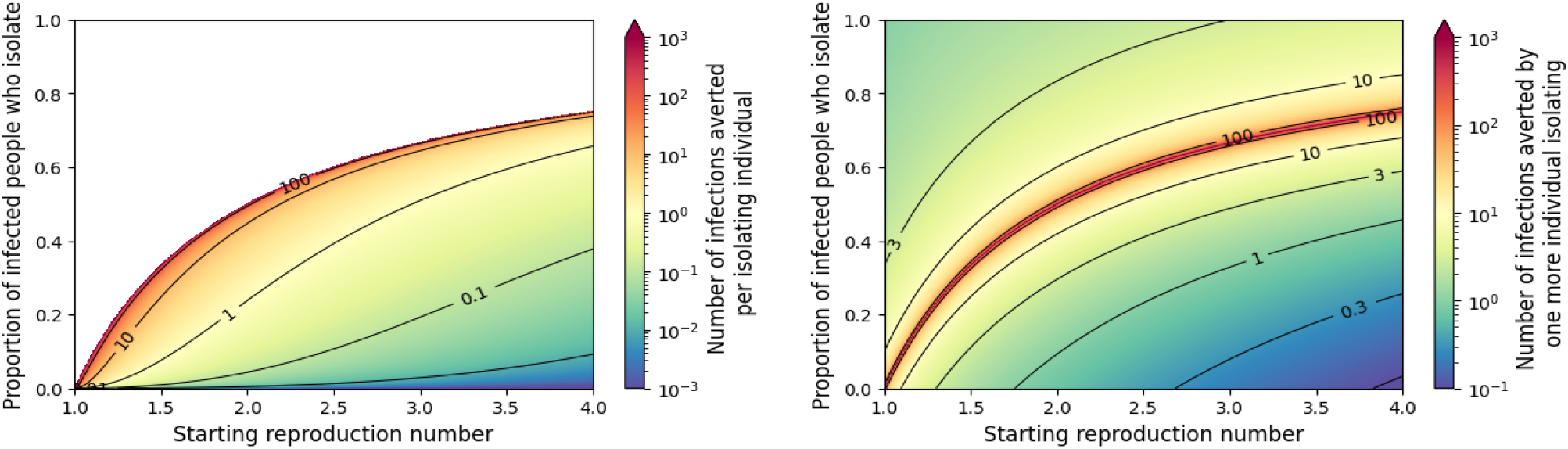
Measuring the effectiveness of many individuals isolating. **Left** The average impact assuming that other interventions set a starting value of R _c_, but an additional fraction of the infected individuals isolate. Calculations are done in the limit of an infinite population. As the fraction of infections isolating increases, the size of the epidemic decreases. Eventually a larger fraction isolating translates to a smaller absolute number isolating due to the smaller epidemic. When enough isolate to drive the resulting R _c_ close to 1 almost all infections are averted while only a small fraction of the total population isolate, so the number of infections averted per isolating individual diverges. Beyond this threshold (white region), epidemics are impossible and the number of infections averted per infected individual is infinite. **Right** The marginal benefit of one more infected individual isolating. That is, given the fraction isolating and the initial R _c_, this gives the number of infections averted if one additional infected individual who would not isolate is successfully identified and isolated.

### Preventing infection

Above we considered individuals acting to prevent onwards transmission only after their infection. This has no direct benefit to the individual. Now we instead consider an individual acting to avoid infection. This protects the individual and prevents onwards transmission. This would require the individual to either receive a vaccine or prophylaxis which prevents infection.

We first consider the impact of a single individual taking proactive actions to prevent her infection. We assume these actions are fully effective. The probability that she would be infected if she did not take those actions is equal to the fraction of the population infected in the epidemic (often called the *attack rate*). Assuming that the population is well-mixed and susceptibility is uniform across the population, the attack rate A depends only on the mean of the offspring distribution, not on any other details of the distribution. It can be calculated from the implicit *final size relation* (*24*–*26*)

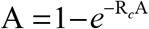

We can think of the attack rate as a function of R _*c*_, that is A= A(R_*c*_).

If an individual makes no behavior change, her probability of being infected is A (R _*c*_). So, if she takes perfectly effective measures to eliminate her chance of being infected, then the direct benefit to herself is a reduction of her infection probability by A (R _*c*_). However, this also prevents transmissions from her to those who would otherwise have been infected by her (her “descendants”). So, if she would have been infected, this reduces the expected number of descendants by an amount F (R_*c*_). The total reduction in expected infections is A(R_*c*_)[1+ F (R_*c*_)]. The first factor A (R _*c*_) represents the probability that the vaccine or other effective disease-avoidance measure prevents her infection. The second factor, 1+ F (R_*c*_) is the expected number of infections that she would have occurred had she been infected, but now do not occur: the 1 represents her own infection, while F (R_*c*_) represents the expected infections averted in her descendants due to the lack of onwards transmission.

Figure 6 tells a surprising story. We consider an individual in the population who avoids infection through vaccine, prophylactic medication, or some other method. When she takes this action, we have no prior knowledge of whether she would be infected or otherwise. The probability her action prevents her own infection is equal to the probability she would have been infected without the action, in other words it is the attack rate A(R_*c*_), shown in the orange curve. So the expected direct benefit to the individual is the probability the individual’s infection is prevented (this can be thought of as the expected number of infections of the individual that are prevented). This increases from 0 at R_*c*_ = 1 to near 1 at R_*c*_ = 4 increases further). (and approaches 1 as R _*c*_

**Figure 6:**
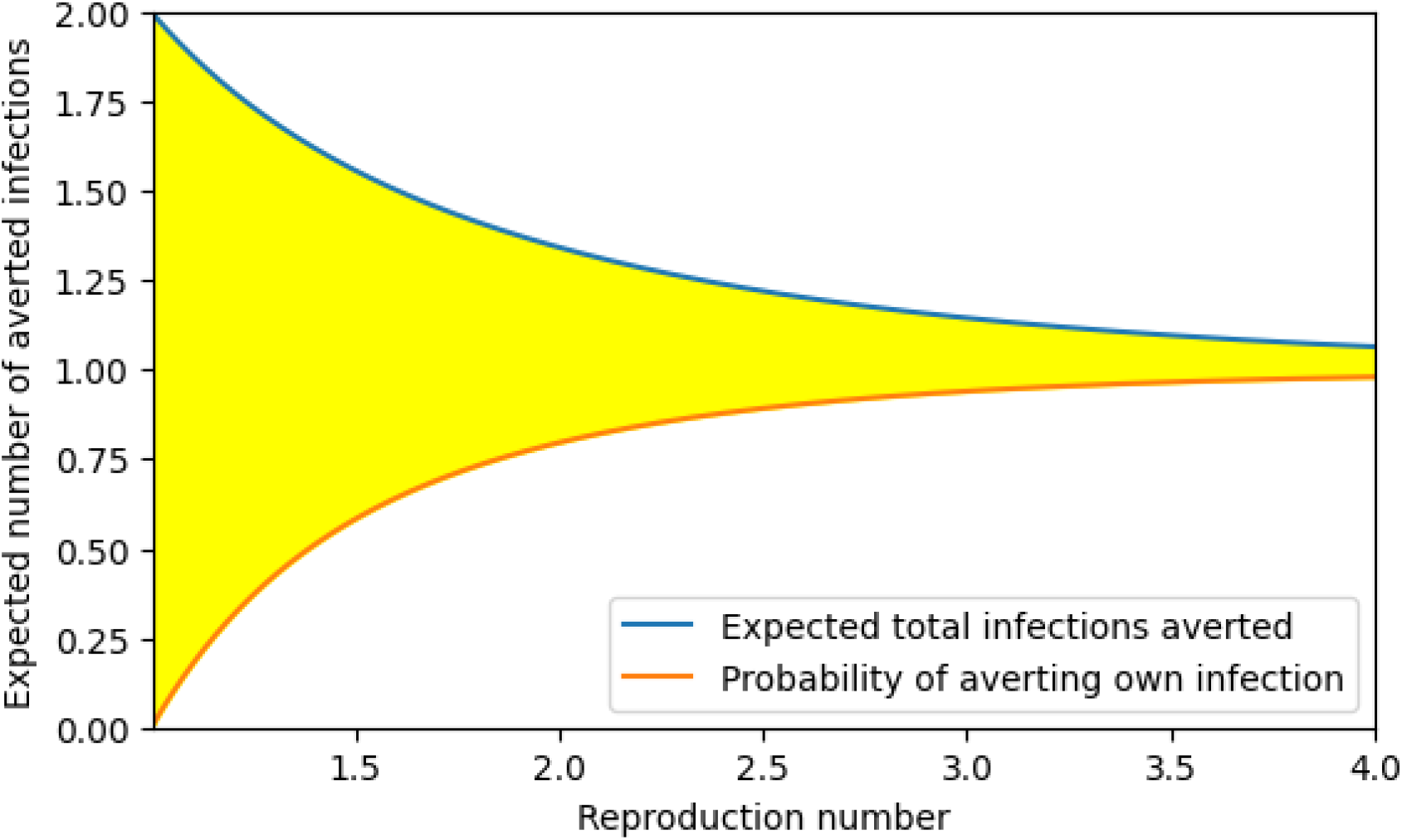
The expected total number of infections averted by an individual taking sufficient action (such as a vaccine or prophylactic medication) to prevent her own infection (top blue curve) for R _c_ > 1. This is partly from the probability that she prevents her own infection [bottom orange curve, equal to A (R _c_)] -- note that the probability of averting her own infected equals the expected number of her own infections averted. The other part of the total benefit is because some of her descendants end up uninfected, shown in the difference between the two curves (shaded yellow) F (R_c_)A(R_c_).

The total number of infections averted includes the possible infection averted of the treated individual as well as the avoidance of infection of some who would have been descendants of this individual. The expected number of infections averted in descendants is the yellow region, and is F (R _*c*_)A (R _*c*_) representing the expected number of infections averted in descendants if she would have otherwise become infected times the probability she would have otherwise become infected. The combined effect (blue curve) shows the expected number of infections averted in the entire population, including the direct effect to the treated individual and the indirect effect to her descendants. Interestingly, the combined effect is always larger than 1 for R _*c*_ > 1.

Because F (R_*c*_) is small for large R _*c*_, due to a high attack rate and significant overdetermination, the expected number of additional infections averted in descendants is very small. So, for large R _*c*_, the expected combined number of infections averted (including her own) is just above 1, with her own benefit constituting almost all of that.

For smaller R _*c*_ approaching 1 however, the probability that the action prevents her infection (A (R _*c*_)) drops and approaches 0. At the same time, the expected number of additional infections averted if infected (F (R _*c*_)) grows to infinity. The combined effect is larger for smaller R _*c*_. For values of R _*c*_ near 1, the expected number of additional infections averted approaches 2, even as the probability that any infections are averted goes to 0. A key additional observation is that for any R_*c*_ > 1, the expected number of infections prevented by a 100% effective vaccine administered is greater than 1.

To make this clearer, consider the example of R_*c*_ = 1.01. The probability a given vaccination directly protects the recipient is small, equal to A≈ 0.0197 (bottom curve of Figure 6). However, in those rare cases in which the recipient does become infected, the number of additional infections comes from a distribution as in Figure 4. From Figure 3, the expected number of additional infections would be F (R _*c*_) ≈ 99.66. The expected number of individuals whose infections are averted due to indirect protection from the vaccine is AF ≈ 1.9670 (shaded part of Figure 6). The combined benefit is about 0.0197 + 1.9670 ≈ 1.9867 (top curve of Figure 6).

In Figure7 we see the impact of multiple individuals taking a vaccine or prophylactic medication or otherwise avoiding infection. A key observation is that as more individuals take self-protective actions, the marginal benefit of the next individual to act increases (left plot), until the epidemic threshold is crossed and epidemics are eliminated. Likewise, the average per-individual benefit increases as more individuals act until the threshold is crossed (right plot). After this, the marginal benefit is 0 and the average impact drops. Remarkably, until the epidemic threshold is reached, each individual that acts prevents (on average) greater than 1 infection.

**Figure 7:**
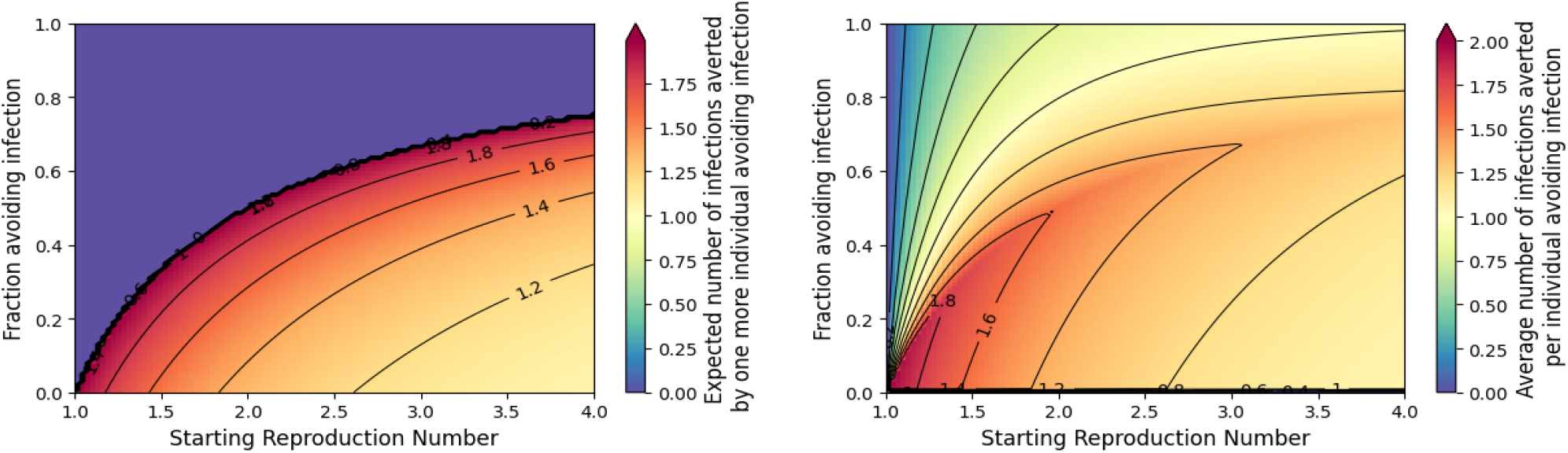
Measuring the impact of many individuals taking actions (such as a vaccine or prophylactic medication) to avoid infection without prior knowledge of whether they would be infected otherwise. **Left**: The marginal reduction in infections from one more individual changing behavior. The marginal benefit increases until the epidemic threshold is reached. Near the epidemic threshold, on average each additional individual acting prevents about 2 infections. Above the critical fraction avoiding infection (purple region), each additional individual has no net impact. **Right**: The number of infections averted over the entire population taking protective action. Until enough individuals take action to prevent an epidemic, the average is greater than 1, and the average increases until the threshold is reached after which it begins to fall.

## Discussion

This is the first paper to model the implications of overdetermination and synergistic collective actions for the outcome which has important implications for evaluating the ethical acceptability of public health measures. We have analyzed the impact of individual and collective behavior modifications related to several important public health measures to control spread of infectious disease, namely quarantine of arrivals, isolation of infected individuals, and the use of vaccines/prophylaxis to prevent infections. A common theme of our results is that there is a synergistic impact: as a larger proportion of the population adopt a protective behavior, the benefit created per individual changing behavior increases.

We now discuss some implications our observations have for ethical policy design.

### Quarantine to prevent epidemic

Border quarantines can prevent the introduction of infection into a population. However, they are costly and rarely perfectly effective. Since our results show that the benefits of quarantine are almost entirely lost if the disease manages to successfully establish itself within the community, the policy cannot allow even one single successful incursion (i.e., an introduction that results in sustained transmission). Our analysis also shows that any single individual’s compliance with quarantine will only have a large impact where there is near perfect compliance amongst others. This is because, if an epidemic will happen anyways, then the impact of an additional introduction from a quarantine breach is generally minimal. The impact of a quarantine leak once disease spread is already established in the population is equivalent to having an infected individual who could isolate failing to isolate, which is discussed below.

It should be noted that if we assume that the community will be able to introduce interventions that would successfully eliminate the introduction, then the balance changes somewhat. In this case the risk of introductions (provided they occur during a period of elimination) is additive, and so each individual who does not comply with the intervention poses a separate risk to the population.

### Isolation to avoid transmission

We find that isolation after infection can be a highly efficient intervention. The high efficiency is for two reasons.

- First, as a larger fraction of the infected individuals isolate, the total number infected drops, which tends to limit the number that need to isolate. Once the *fraction* isolating becomes large enough, the drop in total cases is sufficient that the *number* who isolate also drops.
- Second, the intervention only targets those who are infected, and only when they are infected. This means that those who are never infected are not burdened by the intervention (although they benefit from the compliance of infected individuals).

Isolation is most efficient on a per-individual basis when the reproduction number is close to 1, or if a large enough fraction is already isolating to reduce the reproduction to near 1. Near the epidemic threshold each individual who isolates prevents, on average, the infection of a large number of people (though still small compared to the population size). At the epidemic threshold, the distribution of number of infections prevented is a power-law (a straight line on a logarithmic plot). In these situations, large outbreaks are rare, but they are not so uncommon that we can ignore them – though they are very rare, their very large size means that the average outbreak is large.

Isolation of newly infected individuals requires the ability to quickly identify infected individuals, perhaps through the use of rapid tests or effective contact tracing. In cases where there is asymptomatic or presymptomatic transmission, this will generally be more difficult (*27, 28*)

### Actions taken to avoid individual infection

An individual may avoid infection in several ways. If an effective vaccine or prophylactic medication is available, then he may be able to use these to avoid infection. Alternately, the use of personal protective equipment (PPE), or behavior changes can reduce the probability of infection. If the measures taken are less than 100% effective, the expected impact is scaled by the corresponding factor.

These options to avoid infection may impose costs on an individual even though we do not know in advance whether he would ever be infected without them. This cost can be mitigated in part by changing the level of protection based on local prevalence, *e*.*g*., by wearing PPE only when the risk of infection is high.

A key observation is that for an intervention such as a perfect vaccine, unless the control threshold is crossed and the epidemic is eliminated, every vaccine prevents on average more than one infection. Our analysis allows us to quantify how that benefit is distributed. Most of the benefit goes to the vaccine recipient if R _*c*_ is large. In contrast, if R _*c*_ is near 1, the protective measure has almost no impact on the individual’s probability of infection, but it has a large indirect effect on others in those rare cases where an individual does become infected. This is because overdetermination is reduced in low transmission settings, so that the net expected number of infections averted approaches 2. For the example of R_*c*_ = 1.01 shown in the results section, the probability that vaccine prevents infection of the recipient is about 2%. But when this happens, on average about 100 subsequent infections are prevented. So on average about 2%×100 = 2 infections are averted, but about 99% of the benefit is due to indirect protection of those not vaccinated.

### Ethical and policy implications

Some of our results are relatively unsurprising: if compliance with a quarantine is low, then restricting the liberty of those subject to quarantine is hard to justify if the goal is to prevent an epidemic. However, some of our other results may be more counterintuitive. Specifically, when R _*c*_ is just a little above 1, individual actions have more impact than when R _*c*_ is well above 1 (due to significant overdetermination at high transmission).

In the case of isolating after infection, the expected benefit can be very large if R _*c*_ is close to 1. However, that benefit is enjoyed by someone other than the one enduring isolation. In the case of perfect vaccination/prophylaxis the expected number of infections averted is always at least 1 if R _*c*_ > 1. However, when R _*c*_ is close to 1 the benefit is almost entirely experienced by those not receiving the intervention. These observations provide quantitative data to inform ethical policy design for interventions in cases where the cost of compliance is borne by individuals who receive little or no benefit from compliance.

As a general rule, we see that these interventions are most effective on a per individual basis when the reproduction number is close to 1. When more individuals comply with an intervention, this acts to reduce the overall reproduction number, and the public health benefits per individual affected by the intervention increase – each additional individual complying increases the average effectiveness of those already complying (due to reductions in overdetermination).

Our results speak to the benefit provided by additional individuals complying with policies to isolate/vaccinate/etc., but this requires that the policies exist with resources available so that, for example, an individual can be tested with a high-accuracy test. This may have implications for the level of resources devoted to areas such as providing paid leave for infected individuals, to hiring additional contact tracers, or to the provision of rapid testing to the general population. If the background reproduction number is close to 1, then the benefit of these interventions will be much higher than if it is far from 1.

### Punitive policies are often unjustifiable

Our observations lend support to the argument that for a highly infectious disease, if compliance is not high, then there is little ethical rationale to punish individuals for imposing risks. In high transmission contexts, each individual makes little difference to overall population-level harm beyond their own infection status and outcome (although we are ignoring health system capacity constraints in this analysis). While some might think that punitive or coercive policies might be justified by the goal of raising compliance, it may be more productive to communicate the benefits of collective action.

### Promoting collective action

Population benefits of collective compliance increase with greater compliance. One implication might be that public health messaging should promote high levels of compliance as a good to society rather than stigmatize those who fail to comply. It is perhaps not widely recognized that large numbers of people acting together to reduce transmission of an infectious disease result in *synergistic* public health benefits. To the extent that individuals are aware of such patterns and act on this knowledge, this might reinforce behavior that improves epidemic control through higher compliance.

Our modeling illustrates that the public health consequences directly related to an individual’s noncompliance with interventions is largest when R _*c*_ is near 1, for which overdetermination is rare and the epidemic might seem under control. Communicating the benefits of collective action might help to improve public cooperation with control measures, especially by making people aware that in a highly susceptible population it is all the more important that people (continue to) contribute to control measures as they drive R _*c*_ close to 1, where someone might reasonably conclude his individual incentive to comply is low (as his risk of infection is negligible) (Figure 3 and Figure 6).

### Support for individuals who isolate

A key consequence of our results is that there is a disparity in who receives the benefits from individual actions. Near R _*c*_ = 1 an individual’s action produces much more public benefit than private benefit. This suggests the need for policies based on reciprocity that provide support for individual compliance (e.g., sick leave or isolation payments) are likely to improve the ethical acceptability and effectiveness of public health measures to reduce transmission.

### Limitations and future work

There are a number of limitations to this work that should be addressed in future analysis.

Mathematically, we have studied a relatively simple model of disease – using an SIR model without any incubating or asymptomatic stages and once recovered individuals remain immune. We treat the population as fully mixed, with all individuals equally susceptible and ignore variable risks of severe outcomes. We have focused our attention on measuring the benefit of an intervention – the costs should be explicitly measured as well to balance against those benefits. A more nuanced model will be needed to investigate issues related to more complex disease and population structure. Additionally, our focus has been on basic measures of impact, namely the prevention of an epidemic and reduction in infections. Typically, we expect the number of infections to be a good proxy for morbidity and mortality, with a constant proportion of infections having severe outcomes, though a more realistic model would account for differences within distinct subgroups having different mixing patterns. We have not considered the fact that infected individuals may occupy health care resources that prevent others from accessing care. Nor have we considered issues such as the importance of minimizing the epidemic peak as a separate issue from minimizing total infections.

We have assumed that the background conditions are constant. The interventions are assumed to be unchanging, while in reality they may change in response to the epidemic dynamics. In a real-world scenario we might expect that treatment methods might improve over time; thus a delayed infection might be less severe than an earlier infection. We might also anticipate that a vaccine may be developed, thus a delayed infection may actually end up being a prevented infection or a less-severe infection. On the other hand, we have also assumed that the disease remains unchanged. In reality more transmissible variants may emerge, and so delaying early transmission may come at the cost of a significantly larger later epidemic.

Although this analysis is intended to be widely applicable to diseases which induce immunity, obviously, many readers will focus specifically on the implications for SARS-CoV-2. To avoid it being overinterpreted in that context, we discuss applications and limitations for applying it to the SARS-CoV-2 pandemic. The most significant limitation is that our model does not include the role of multiple variants. The variants have driven much of the later phases of the pandemic through avoiding immune response. Thus, our results are most appropriately applied to just a single wave. Once the Delta variant was replaced by the Omicron variant, the effectiveness of the original monovalent vaccine against transmission dropped significantly (*29*). Inasmuch as the indirect benefit to others from vaccine blocking transmission is reduced, our results about the benefit of effective prophylactic measures no longer apply to the original monovalent vaccines.

Our results showing that the benefit of a single individual’s actions drop as R _*c*_ rises remain valid for each later wave, either because of changes in the background interventions or changes in the infectiousness of the virus. Our modeling results imply that the ethical implications of an individual’s behavior are significantly reduced by overdetermination in waves which have higher R _*c*_ whether this is because the disease might be more transmissible or because a significant proportion of other individuals are not taking protective measures.

## Materials and Methods

In this section we derive our mathematical approaches and perform some of the technical analysis of the model. We begin by briefly describing the model assumptions and providing two small examples demonstrating key features of stochastic infection spread. We then derive the mathematical approaches. Finally, we perform some of the analysis, ending with a rigorous derivation of the observation that as R_*c*_ →1^+^ completely effective vaccine approaches 2. the expected number of infections prevented by a

### Model Assumptions

We assume a stochastic model of infection spread, based on the standard Susceptible-Infected-Recovered (SIR) model (*30, 31*). Each infected individual potentially transmits to *k* others, chosen uniformly at random from the population, where *k* is chosen from some prescribed distribution. If the recipient of a transmission is susceptible, an infection event occurs. If not, the transmission has no effect. The distribution of the number of transmissions *k* is known as the *offspring distribution*, with *p*_*k*_ denoting the probability of *k* transmissions. After transmitting to its *offspring* the infected individual recovers with immunity.

If there are no interventions in place, then the average of the offspring distribution is the *basic reproduction number* R _0_. If interventions are in place, then the average of the offspring distribution after accounting for those interventions is the *reproduction number under control* R _*c*_. We assume that R _*c*_ remains constant for the duration of the epidemic. Because transmissions have no effect when the recipient has already been infected, we also introduce the *effective reproduction number* R_eff_ (*t*) which measures the average number of successful transmissions from an infected individual. R_eff_ accounts for both the intervention and the immunity that the population has developed. We use R_∞_ to denote R _eff_ (∞). It is less than 1 and equals R_*c*_ times the proportion of the population that is susceptible at the end of the epidemic 1− A (R _*c*_).

It is known that the probability an epidemic becomes established is sensitive to the frequency of superspreading, the tendency that a small fraction of the infected individuals cause a large fraction of the transmissions (*19, 21*–*23*). To investigate the significance of superspreading in our analysis, we consider the impact of a Poisson offspring distribution and a negative binomial distribution. The Poisson distribution is parametrized by a single variable, the mean R _*c*_, and individuals who transmit significantly more than average are negligibly rare. The negative binomial distribution is parametrized by the mean R _*c*_ and a dispersion parameter, which we take to be 0.16, which includes significant superspreading. This is consistent with estimates for SARS (*22*) and allows us to clearly show what impact (if any) superspreading has. More recent estimates for SARS-CoV-2 suggest that the dispersion parameter may be larger (*21*); however our goal is to investigate the qualitative impact of superspreading rather than making exact predictions for a particular disease.

### Example Outbreaks

In Figure 8, we show transmission chains in two outbreaks in populations of 50 individuals. The two populations have R_*c*_ =1.5 and R_*c*_ = 2.5. Each infected individual transmits independently to each of the other 49 individuals with probability R_*c*_ / 49, resulting in a distribution that is approximately Poisson with mean R_*c*_. In both cases, the outbreak successfully establishes and only terminates because eventually a significant number of transmissions go to previously infected individuals. In a larger population, those blocked transmissions would have gone to other susceptible individuals, resulting in a large-scale outbreak, that is, an *epidemic*.

**Figure 8.**
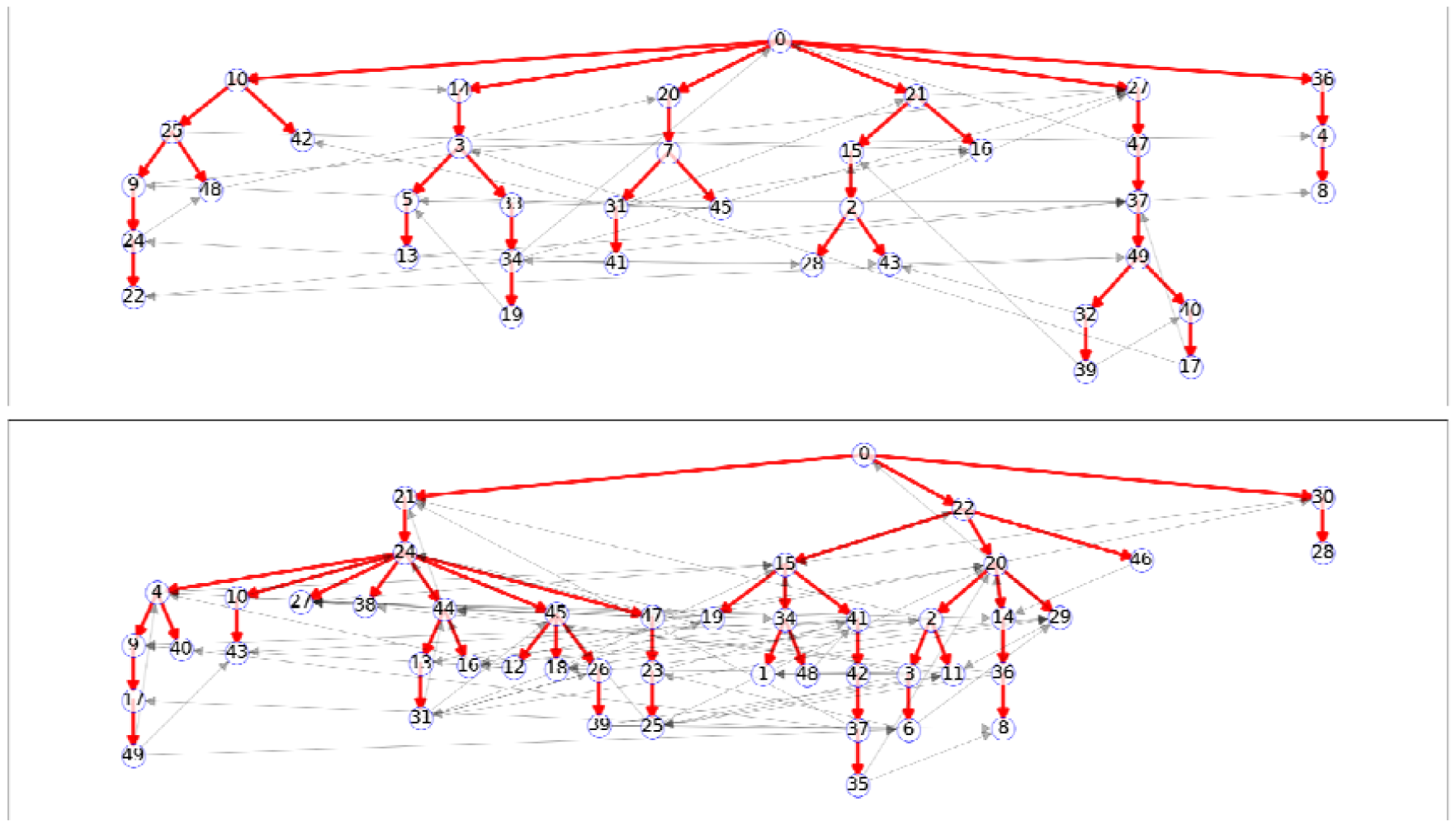
Two sample outbreaks with R_c_ =1.5 (top) and R_c_ = 2.5 (bottom), each starting from a single infection in a population of 50 individuals. Only the eventually infected nodes are shown. The red lines denote successful transmissions while the gray lines denote transmissions from an infectious individual to one who had already been infected. Note that given the transmissions in the R_c_ =1.5 case, had individual 36 (second row on right) been the initial infection, only individuals 36, 4, and 8 would have been infected. Additionally, had individual 49 (fifth row on right) isolated after infection (or been effectively protected from infection), this would have prevented the infections of 32, 39, 40, and 17. In the R_c_ = 2.5 case, there are many more (potential) transmission chains: more infections occur, transmission chains tend to be longer, and removing one individual tends to protect fewer others.

Figure 8 shows that although many epidemics spread far and are limited only by the population size, outbreaks starting from some individuals would not spread far. For example, in the top plot if the infection introduced in individual 42(second node in third level) rather than individual 0 or if in the bottom plot infection started with 28 (last node in third level), these would not lead to long transmission chains. However, many of the other individuals would spark large-scale transmission through the population. So, if multiple introductions occur, it is likely that more than one of them is sufficient to spark an epidemic. This is more likely for larger R_*c*_. This illustrates how *epidemics* can be overdetermined, which plays an important role in our analysis.

Additionally, we see that individual *cases of infection* may or may not be overdetermined. If we blocked transmission from some individuals it would provide effective protection to others. For example, in the top plot preventing transmissions from individual 49 (lower right) would be sufficient to prevent the infection of 32, 39, 40, and 17. However, many infections are overdetermined because there are alternate transmission routes. For example, in the top plot preventing transmissions from 10 (first node in second level) would prevent the infection of 25, but all other descendants would eventually be infected through alternate chains of transmission. The existence of multiple transmission chains to the same individual becomes more likely for larger R_*c*_.

In a practical setting we do not know *a priori* which transmissions would occur. Based on the offspring distribution, we can calculate the probability that an introduced infection results in an epidemic. We can also calculate the distribution of the number of infections averted by one individual’s behavior change. However, due to the stochasticity inherent in the system, for a specific infected individual introduced to a population, we cannot know in advance whether he would spark an epidemic, or in an ongoing epidemic, we do not know exactly how many infections another individual might avert by changing her behavior. Thus, our analysis will focus on the expected (*i*.*e*., the average) impact over many realizations.

The key quantities we focus on are:

- The expected impact of a single infected individual acting alone or multiple infected individuals acting together to isolate prior to entering a community to prevent an epidemic.
- The expected impact of a single individual acting alone or multiple individuals acting together to reduce the total number of infections occurring in an epidemic.

### Mathematical Methods

We now build up the mathematical methods used to analyze the sort of outbreaks that can occur. We will assume throughout that the population size is very large. Under this assumption two typical outcomes occur in large populations: either an outbreak remains small and dies out quickly or it becomes an epidemic that grows until it is limited by the population size. If R_*c*_ <1 only small outbreaks occur. If R_*c*_ >1 large epidemics can occur, but small outbreaks are still possible. The size distribution of the *number* infected in small outbreaks is independent of the population size but does depend on the offspring distribution. In contrast, in a large epidemic the *proportion* infected is independent of the population size (so the number infected is proportional to the population size) and the proportion is independent of the offspring distribution.

### Probability Generating Functions

In this section we briefly introduce some properties of probability generating functions (PGFs). More complete details can be found in (*23, 32*).

The number of outgoing edges from a particular node is chosen from the offspring distribution. If *p*_0_, *p*_1_, … represent the probability of zero, one, … offspring, then the PGF of the distribution (*32*) is defined to be

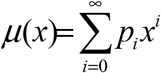

We have considered a Poisson distribution with mean R_*c*_ for which *μ*(*x*)= exp(−R_*c*_(1− *x*)) and a negative binomial distribution with a dispersion parameter of *κ* = 0.16 for which 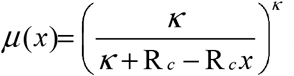, but any probability distribution can be chosen based on the characteristics of the disease. The mean of the offspring distribution satisfies 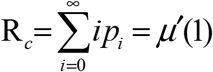.

In many cases it may be difficult to find the coefficients of a PGF *μ*(*x*) directly, but it is possible to calculate the values of *μ*(*x*) to high precision on the unit circle in the complex plane. Then we can use a Cauchy integral to find arbitrary coefficients of *μ*(*x*) [see section A.1 of (*23*)] (in fact if we parametrize the unit circle by the angle *θ*, then *μ*(*x*) becomes a complex-valued Fourier Series in *θ*, and the Cauchy integral becomes the formula for the coefficients of a Fourier Series). In the context of disease spread, if Ω_∞_ (*x*) is the PGF for the final size distribution when the reproduction number is less than 1, using approaches shown below, it is possible to calculate Ω_∞_ (*x*) at arbitrary values of *x*. Then this approach can be used to find the coefficients of the series expansion of Ω_∞_ (*x*) (*23*).

### The final size relation

We have assumed homogeneous susceptibility in a well-mixed large population. It is well-known that the final size of epidemics under these assumptions depends on R _*c*_, the average of the offspring distribution, but it does not depend on finer details of the offspring distribution. Here we briefly derive the final size relation following (*24, 25*) [see also (*26*) for related results]. We let A(R_*c*_) denote the expected proportion of the population infected in an epidemic. Taking *N* to be the (large) population size, the total number of infections is A (R _*c*_)*N*. Since on average they each produce R _*c*_ transmissions, the total number of transmissions that occur in the epidemic is well-approximated by R_*c*_ ·A(R_*c*_)· *N*. On average each member of the population thus receives R_*c*_ ·A(R_*c*_) transmissions. In a large well-mixed population, we can reasonably assume independence of these events. This means that the number of transmissions received is Poisson-distributed with mean R_*c*_A(R_*c*_). The probability of receiving no transmissions is 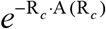. However, because this is the probability of not being infected it also equals the proportion of the population that remains susceptible. We arrive at the final size relation

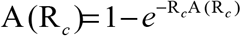

This can be solved iteratively by setting 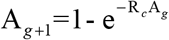 with A_0_ =1.

### Calculating epidemic probability

To calculate the epidemic probability, we consider a process known as the Galton-Watson process (or birth-death process). Let *α* denote the probability that a given individual in a branching process has a finite number of descendants. Then 1−*α* is the probability of an infinite number of descendants. The number of descendants is finite exactly when every single offspring has a finite number of descendants. Since each offspring has a finite number of descendants also with probability *α* we find

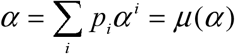

Thus, the probability of an infinite number of descendants can be calculated by finding the roots of *α* = *μ*(*α*) where *μ*(*x*) is the PGF. One root is always *α* =1 corresponding to never having an infinite number of descendants, but if there is another root, it will lie between 0 and 1, and it is the correct root to choose. This other root exists when R _*c*_ > 1.

In practice we can find *α* by setting *α*_0_ = 0 and letting *α*_*g* +1_ = *μ*(*α*_*g*_), until the values found for *α* _*g*_ converge. In this approach, *α* _*g*_ can be interpreted as the probability that the outbreak terminates by generation *g*.

### Calculating the impact of isolation after infection

To determine the expected impact of isolation of *u* after becoming infected, we need to calculate the expected number of descendants an individual who would not be infected through some transmission path not through *u*, as shown in Figure 9. To determine this, we consider the *residual offspring distribution*, that is the distribution of the number of direct offspring who would not be reachable along any other transmission chain. If an individual would cause *i* transmissions, a fraction A(R_*c*_) of them go to individuals who would otherwise be infected through a different chain, as would all their descendants, and so only a fraction 1− A (R _*c*_) are successful.

**Figure 9:**
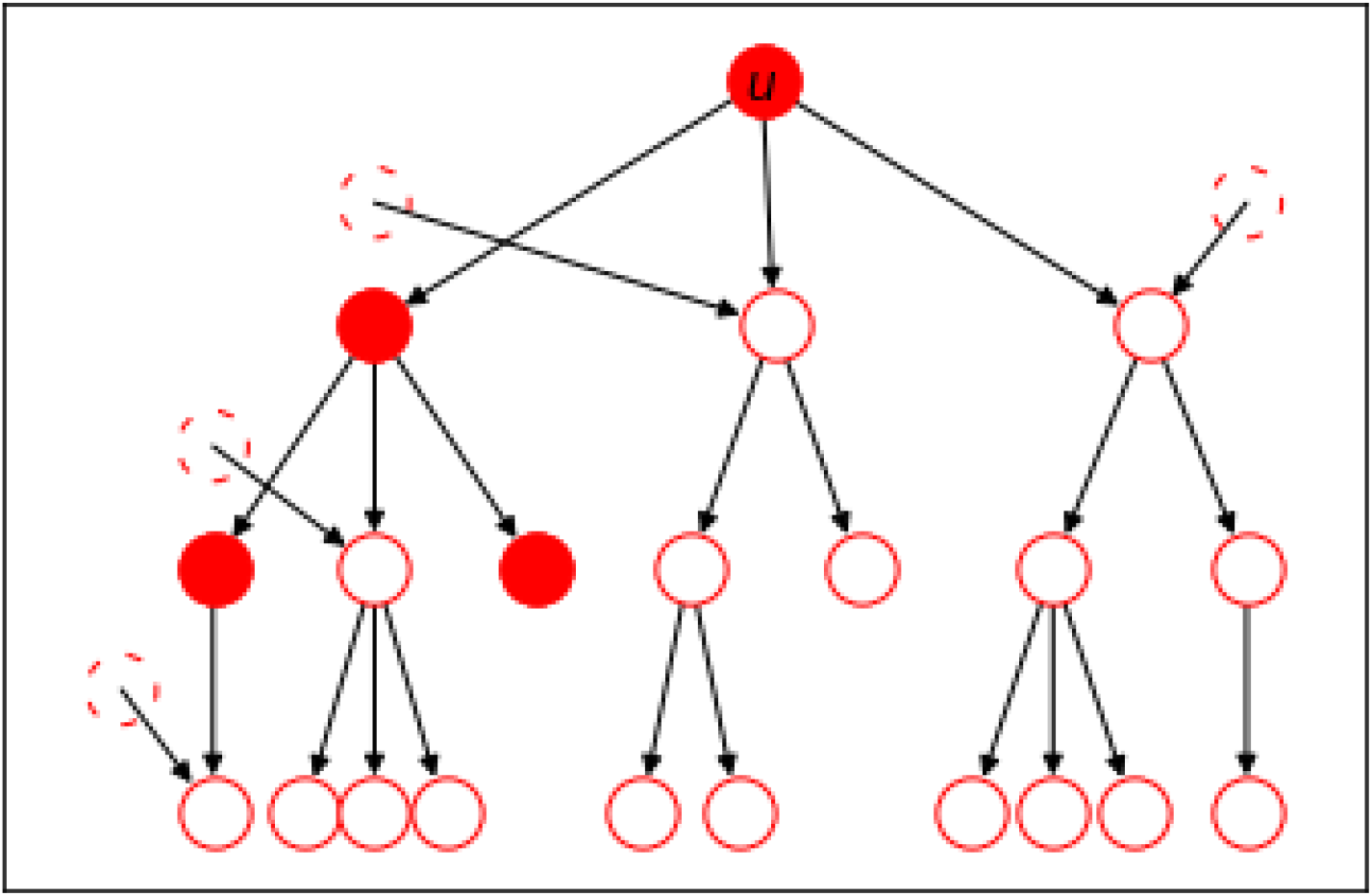
A schematic showing the descendants of u with filled circles showing those whose infections would be averted if u isolates after infection. Dashed hollow circles denotes infected individuals that are not descendants of u who provide additional transmission paths to some descendants of u. Hollow circles with solid boundaries denote the individuals whose infection is overdetermined.

The average of the residual offspring distribution is R_∞_ =[1−A(R_*c*_)]R_*c*_, which is the initial reproduction number multiplied by the fraction who remain susceptible at the end. This is the “effective reproduction number” at the end of the epidemic. If R_*c*_ ≤1, then A(R_*c*_) = 0 and we find R_∞_ = R_*c*_. However, if R_*c*_ >1, then 0 < A (R _*c*_) < 1, and it turns out that R_∞_ is less than 1 (otherwise the epidemic would never stop growing). The expected number of infections averted among those who are reachable from a path of *g* generations from *u* ends up being 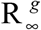. Summing this over all *g*, we find that the initial infected individual’s isolation prevents

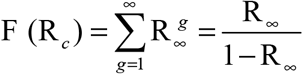

infections, where R_∞_ =[1−A(R_*c*_)]R_*c*_ is the effective reproduction number at the end of transmission in a population having reproduction number under control of R_*c*_. Note that when R_∞_ is close to 1, this is large. This happens when R_*c*_ is close to 1.

To go further, we can calculate the distribution of the total number of infections averted. The PGF for the residual distribution is *μ*(R_∞_*x* +1-R_∞_). Following methods derived in (*23*), the PGF for the size distribution of the number of infections averted is Ω_∞_(*x*)= *xμ*(1− R_∞_ +R_∞_Ω_∞_(*x*)).

This can be calculated for arbitrary values of *x* by iterating Ω_*g*+1_(*x*) = *xμ*(1− R_∞_+R_∞_Ω_*g*_ (*x*)), starting with Ω_0_(*x*) =1. Iterating for values of *x* on the complex unit circle until the results converge gives us Ω_∞_ (*x*) at those values. Numerically approximating a Cauchy integral, this allows us to calculate the individual coefficients of Ω_∞_ (*x*), from which we know the distribution of the total number of infections averted.

To calculate the impact of multiple individuals isolating, we note that if a fraction *ρ* of the population isolate after infection, then the offspring distribution is modified. With probability *ρ* an infected individual isolates and causes no infections, while with probability 1− *ρ* they cause a number of infections chosen from the original distribution. This means that R_*c*_ is effectively multiplied by 1− *ρ*. We can redo the calculations for final size and individual impact above using (1− *ρ*)R_*c*_ instead of R_*c*_.

### Calculating the impact of avoiding infection

If an individual either gets a 100% effective vaccine or prophylactic medication, she can avoid infection. Doing so, she prevents her own infection with probability A(R_*c*_), as it has no effect if she would have avoided infection anyways. When she prevents her own infection, the total number of infections averted due to this is 1+F (R_*c*_) where the 1 accounts for protection of herself and F (R_*c*_) accounts for protection of others. Combining this reduction with the probability that the reduction occurs, we find that the expected reduction in infections due to a single individual taking measures to prevent her own infection is A(R_*c*_)[1+ F (R_*c*_)].

To calculate the impact of multiple individuals getting vaccinated or otherwise avoiding infection, we note that if a fraction of the *ρ* population is immune to infection, then the offspring distribution is modified. With probability *ρ* a random transmission is blocked by the protection to the recipient. This means that R_*c*_ is effectively multiplied by 1− *ρ*, though for a different reason than before. We can redo the calculations for final size and individual impact above using (1− *ρ*)R_*c*_ instead of R_*c*_.

We now derive the expected number of infections averted for the limit approaching R_*c*_ =1 from above, showing that it matches the apparent limit of 2 shown by the numerics. We need to calculate 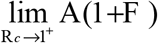 where 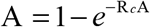 and F = R_∞_ / (1− R_∞_) = [1− A]R _*c*_ / (1−[1− A] R _*c*_).

To find this value, we invert the relationship between A and R _*c*_ (for R _*c*_ > 1), writing 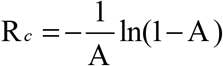. We now take the limit A

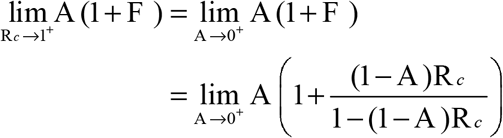

Substituting for 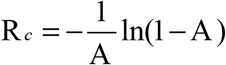, we get

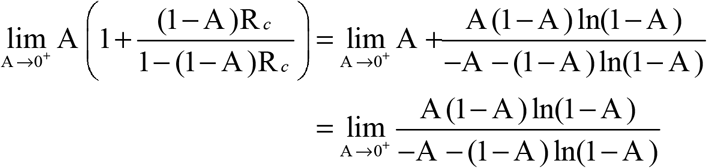

Taking one round of L’Hopital’s rule yields

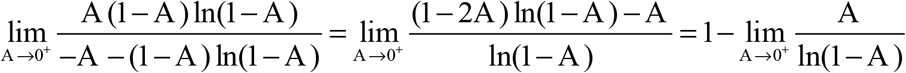

Applying L’Hopital’s rule again yields

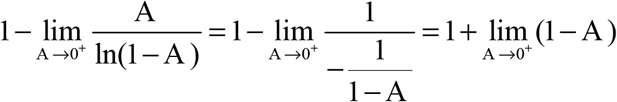

which is 2.

### Extensions to multigroup populations

It is natural to consider a population with multiple subgroups having different mixing patterns and different disease impacts (e.g., this might be multiple age classes). Although we do not attempt that here, the generalization of the methods we have used is straightforward. The application of probability generating functions to a multi-type branching process is described in (*23*). This would allow us to address the probability of an epidemic when quarantine is considered.

To analyze the expected impact an individual has on the number of infections, we would first calculate the final size of the epidemic in each subgroup, as described in (*24*). Then we would need to generalize R_∞_ to a matrix that captures the transmissions between and within groups at the end of the epidemic. This would allow us to calculate the expected number of descendants accounting for overdetermination, that is, through the residual population after allowing the rest of the epidemic to spread.

Finally to find the distribution of the number of descendants (after accounting for overdetermination), we would again use the final size of the epidemic in each subgroup, and then we would follow (*23*) to find a distribution of the multi-type branching process through the residual population.

## Data Availability

All data presented are based on mathematical calculations. The code to produce the figures is available at https://github.com/Joel-Miller-Lab/SIR-modeling-for-ethical-interventions

## Funding

- Australian Mathematical Sciences Institute summer research fellowship (DR, JCM).
- CaRE grant scheme from La Trobe University’s School of Engineering and Mathematical Sciences (DR, JCM)
- The Wellcome Trust [203132] and [221719] (EJ)

## Author contributions

Conceptualization: DR, GH, JCM, MS, EJ

Methodology: JCM & DR

Investigation: DR & JCM

Figure Generation: DR & JCM

Writing—original draft: DR & JCM

Writing—review & editing: EJ, DR, GH, MS, JCM

Rigorous derivation of limits: ACS

## Competing interests

Authors declare that they have no competing interests

## Data and materials availability

All data are contained within the paper. Code necessary to create the figures can be found at https://github.com/Joel-Miller-Lab/SIR-modeling-for-ethical-interventions

## Notes

### Competing Interest Statement

The authors have declared no competing interest.

### Summary of Updates

This version has been updated in response to reviewer comments.

